# Effects of catheter-based renal denervation in hypertension: a systematic review and meta-analysis

**DOI:** 10.1101/2024.06.12.24308869

**Authors:** Davor Vukadinović, Lucas Lauder, David E. Kandzari, Deepak L. Bhatt, Ajay Kirtane, Elazer R. Edelman, Roland E. Schmieder, Michel Azizi, Michael Böhm, Felix Mahfoud

**Author notes:** Corresponding author: Davor Vukadinović, MD, Saarland University Hospital Klinik für Innere Medizin III, Kardiologie, Angiologie und Internistische Intensivmedizin Kirrberger Strasse 100, D-66421 Homburg, Germany; / 16-15500, / 16-15995.

## Abstract

**Background:** Several sham-controlled trials have investigated the efficacy and safety of catheter-based renal denervation (RDN) with mixed outcomes.

**Aim:** To perform a comprehensive meta-analysis of all randomized, sham-controlled trials investigating RDN with first- and second-generation devices in hypertension.

**Methods:** We searched MEDLINE and Cochrane Library for eligible trials. Outcomes included both efficacy (24-hour and office systolic [SBP] and diastolic blood pressure [DBP]) and safety (all-cause death, vascular complication, renal artery stenosis >70%, hypertensive crisis) of RDN. We performed a study-level, pairwise, random-effects meta-analysis of the summary data.

**Results:** Ten trials comprising 2,478 patients with hypertension while being either off- or on-treatment were included. Compared with sham, RDN reduced 24-hour and office systolic BP by 4.4 mmHg (95%CI −6.1, −2.7, p<0.00001) and 6.6 mmHg (95%CI −9.7, −3.6, p<0.0001), respectively. The 24-hour and office diastolic BP paralleled these findings (−2.6 mmHg, 95%CI - 3.6, −1.5, p<0.00001; −3.5 mmHg, 95%CI −5.4, −1.6, p=0.0003). There was no difference in 24-hour and office SBP reduction between trials with and without concomitant antihypertensive medication (p for interaction 0.62 and 0.73, respectively). There was no relevant difference concerning vascular complications (OR 1.69, 95%CI 0.57-5.0, p=0.34), renal artery stenosis (OR 1.50, 95%CI 0.06-36.97, p=0.80), hypertensive crisis (OR 0.65, 95%CI 0.30-1.38, p=0.26) and all-cause death (OR 1.76, 95%CI 0.34-9.20, p=0.50) between RDN and sham groups. Change of renal function based on eGFR was comparable between groups (p for interaction 0.84). There was significant heterogeneity between trials.

**Conclusions:** RDN safely reduces ambulatory and office SBP/DBP vs. a sham procedure in the presence and absence of antihypertensive medication.

**Clinical Perspective:** What is new?

- Several sham-controlled trials have investigated the efficacy and safety of catheter-based renal denervation (RDN) with mixed outcomes.
- This comprehensive meta-analysis comprising 2,478 patients shows that irrespective of the utilized method (radiofrequency-, ultrasound-or alcohol-mediated), renal denervation effectively reduced ambulatory and office systolic blood pressure.
- Renal denervation exhibited no additional risk concerning vascular injury or renal function impairment.

What are the clinical implications?

- This meta-analysis supports current guidelines/consensus statements that renal denervation represents an additive treatment option in carefully selected patients with uncontrolled hypertension.

## Introduction

Renal denervation (RDN) reduces blood pressure (BP) by modulating efferent and afferent sympathetic renal nerve activity. ^1^ Although early unblinded studies suggested a pronounced BP-lowering effect of RDN in patients with severe treatment-resistant hypertension ^2^, the first generation of sham-controlled randomized trials ^3–5^ was unable to prove the BP-lowering efficacy of radiofrequency (RF) RDN using a mono-electrode catheter compared with a sham procedure. Several procedural and methodological confounders were identified and subsequently addressed by well-designed second-generation sham-controlled trials. ^6–11^ Moreover, human cadaveric anatomical studies provided important insights into renal sympathetic nerve distribution and density that informed the design of revised procedural techniques and technologies. ^12, 13^ These second-generation clinical trials utilized refined catheter systems, such as multi-electrode RF, ultrasound, and alcohol-mediated RDN, in patients with mild-to-moderate hypertension in the presence and absence of antihypertensive medications to resistant hypertension despite single-pill triple agent combination therapy. ^14–18^ Previous meta-analyses of sham-controlled trials investigating the Spyral and Paradise RDN catheter systems ^19^ and a subgroup analysis (with sham-controlled trials) of another meta-analysis ^20^ consistently showed that RDN reduced office and 24-hour SBP. We performed an updated comprehensive meta-analysis on summary data from all published randomized, sham-controlled clinical trials investigating the effects of RDN on BP across different devices.

## Methods

### Eligibility criteria, literature search, and study selection

A meta-analysis of the published randomized, sham-controlled trials (RCTs) was performed in line with the Preferred Reporting Items for Systematic Review and Meta-Analyses (PRISMA) statement. ^21^ The protocol for this analysis has been registered at PROSPERO (CRD42022376504). Eligible publications were searched in MEDLINE and Cochrane Library from 01/2000 to 01/2024 using the following keywords and Medical Subject Headings (MeSH) terms: [renal denervation OR renal sympathetic denervation AND randomized sham-controlled clinical trial OR sham-controlled AND treatment arterial hypertension OR treatment hypertension] (Supplement, Table 1 and 2). Furthermore, we had access to several randomized, yet unpublished trials (Netrod RDN ^22^, Iberis HTN ^23^, TARGET BP I ^24^) that have been released at scientific meetings and were submitted for publication. These trials met the criteria for being considered high-quality trials; therefore, we made the decision to include them in the present analysis.

All randomized, sham-controlled trials investigating catheter-based RDN using various methods of nerve ablation (radiofrequency, ultrasound, alcohol-mediated) in patients with hypertension only were considered eligible. Two authors (DV and LL) reviewed the full texts of potentially eligible studies, thereby cross-checking the presence of inclusion criteria. A reference manager software (Zotero, Corporation for Digital Scholarship, Vienna, VA, USA) was used to remove duplicate.

### Data extraction

Two authors (DV, LL) extracted data of interest using a predefined template and assessed risk of bias at study level using the revised Cochrane risk-of-bias tool (RoB 2.0) ^25^, under final supervision of a third investigator (FM). For each included study we assessed the risk of bias for the following domains: randomization process, deviation from intended intervention, reporting of missing data, measurement of outcome, selection of the reported results, single or multicenter trial. Each domain was judged as low, moderate or high risk of bias based on the results of the assessment that were stated as support of judgement. To assess small-study effects, we planned to use funnel plots if at least 10 trials were available. If asymmetry in the funnel plot was detected, we planned to review the characteristics of the trials to assess whether the asymmetry was likely due to publication bias or other factors such as methodological or clinical heterogeneity of the trials.

The following data were extracted: i) baseline characteristics, study design, primary outcome, duration of follow-up, sample size, utilized method of RDN and RDN device; ii) mean change from baseline to primary endpoint assessment in office and ambulatory systolic and diastolic BP (SBP/DBP), daytime and nighttime SBP/DBP, number of antihypertensive drugs at randomization and at the time of primary endpoint assessment; iii) safety data, including the number of events of all-cause death, vascular complication, renal artery stenosis >70%, hypertensive crisis, stroke, hospitalization for heart failure (HF), embolic event, new end-stage-renal-disease (ESRD) and mean change from baseline to primary endpoint assessment renal function according to estimated glomerular filtration rate (eGFR by CKD-EPI).

### Outcomes of interest

The main efficacy outcomes were between-group (RDN vs sham) difference concerning changes from baseline in mean 24-hour SBP and DBP as well as office SBP and DBP. Additional efficacy outcomes were changes from baseline in daytime and night-time SBP/DBP. Safety outcomes included the between-group difference in incidence of previously mentioned safety events and changes in eGFR. Furthermore, we compared the number of antihypertensive drugs at the time of primary endpoint assessment between the groups, to assess whether RDN may impact medication burden. Also, we looked for the number of patients who met escape criteria and experienced antihypertensive medication adjustments (addition/cessation of new drugs or changes in drug doses) due to very high or low BP values during or after the follow-up in the included trials.

### Statistical analysis

A study-level, pairwise, random-effects meta-analysis was performed based on the intention-to-treat or modified intention-to-treat analysis (depending on how data were reported) of the summary data. For efficacy outcomes, we pooled the mean changes of BP with its corresponding SD at the time of primary outcome assessment and applied the inverse variance statistical method. If SD was not reported in the trials, we used the reported 95% confidence intervals (CI) or the standard error of the mean (SEM) to calculate SD. For safety outcomes, we determined odds ratios (ORs) by applying the Mantel-Haenszel method. In case the outcomes had documented null events in one arm (RDN or sham), a fixed value (typically 0.5) would be added (routinely performed by software included in RevMan) to avoid computational error. ^26^ Heterogeneity between the trials was assessed using Cochran’s Q test and I^2^ statistic. Relevant statistical heterogeneity was assumed if I^2^ >50%. In case of relevant heterogeneity for main outcomes, jackknife sensitivity analyses, such as running the analysis after excluding each trial in turn would have been performed to identify outliers. To increase the reliability of the summary findings, we only included high-quality trials according to our assessment and in accordance with the European Society of Cardiology (ESC) Council on Hypertension and the European Association of Percutaneous Cardiovascular Interventions (EAPCI) clinical consensus statement in the main analyses. ^6^ As part of sensitivity analyses, all eligible trials were included for which main (change of ambulatory and office SBP and DBP) and additional efficacy outcomes like change of day- and nighttime SBP/DBP were analyzed. To evaluate the long-term efficacy of RDN, we pooled the longest available follow-up 24-hour and office SBP data.

Further subgroup analyses were conducted to investigate the potential moderator effects of certain factors on the ultimate outcomes regarding the primary efficacy measures. These included only high-quality trials as defined by the ESC/EAPCI clinical consensus document. We determined p for interaction/heterogeneity ^27^ concerning effects of RDN on ambulatory and office systolic/diastolic BP between trials in the following settings: effects of RDN in the presence and absence of antihypertensive medication (i.e., ON- and OFF-medication trials design), first-generation RF device vs second-generation devices, and RDN modality (i.e. radio-frequency vs ultrasound ablation vs alcohol-mediated techniques). Study-specific and summary effect estimates with corresponding 95% confidence intervals (CIs) and p-values were visualized using forest plots. All statistical analyses were performed using RevMan Version 5.4 (The Cochrane Collaboration, Copenhagen, Denmark) and GraphPad Prism Version 6 (GraphPad Software, Boston, Massachusetts, USA). All p-values were two-sided, with p less than 0.05 considered significant.

## Results

The search identified 145 records (Supplement, Figure 1), of which 13 trials were considered eligible. Ten trials ^3, 16–18, 22–24, 28–30^ comprising 2,478 patients, were graded as high-quality trials (Supplement, Table 3-15) (Table 1). Three additional randomized, sham-controlled trials ^4, 5, 31^ graded as moderate risk of bias ^6^ were added to the main analysis and analyzed as a part of sensitivity analysis comprising 2,690 patients. Long-term data on 24-hour and office SBP were available for 5 trials. ^32–34^ The baseline characteristics of all trials are summarized in Table 1. Small study effects have been evaluated using funnel plots for following outcomes: 24-hour SBP/DBP and office SBP/DBP. ^35^ The median follow-up duration to primary endpoint assessment was 3 months, ranging from 2 to 6 months across trials.

**Table 1.** Baseline characteristics of the included trials.

| Study and publication year | Study design | Sample size (RDN/Sham) | Inclusion criteria | Exclusion criteria | Primary efficacy endpoint | Catheter (Modality) | Link to the public trial registry |
| --- | --- | --- | --- | --- | --- | --- | --- |
| SYMPPLICITY HTN-3 2014 <sup>3</sup> | Randomized 2:1, RDN vs. sham procedure, on med | 364/171 | 18-80 y, severe resistant hypertension, office SBP >160 mmHg on $\geq 3$ med | Known secondary hypertension, >1 hypertensive crisis, unsuitable renal artery anatomy, eGFR <45 ml/min/1.73 m<sup>2</sup> | Change in office SBP after 6 months | Symplcity Flex (radiofrequency) | NCT01418261 |
| Desch et al. 2014 <sup>5</sup> | Randomized 1:1, RDN vs. sham procedure, on med | 35/36 | 18-75 y, resistant hypertension, daytime BP 135-149/90-94 mmHg on $\geq 3$ med | Severe comorbidities, unsuitable renal artery anatomy, eGFR <45 ml/min/1.73 m<sup>2</sup> | Change in 24-h SBP after 6 months | Symplcity Flex (radiofrequency) | NCT01656096 |
| ReSET 2016 <sup>4</sup> | Randomized 1:1, RDN vs. sham procedure, on | 36/33 | 30-70 y, resistant hypertension, daytime SBP | Secondary hypertension, congestive HF, unsuitable | Change in daytime ambulatory SBP after 3 | Symplcity Flex (radiofrequency) | NCT01762488 |
| | med | | >145 mmHg on $\geq 3$ med | renal artery anatomy, eGFR $\leq 30$ ml/min/1.73 m<sup>2</sup> | months | | |
| RADIANCE-HTN SOLO 2018 <sup>17</sup> | Randomized 1:1, RDN vs. sham procedure, off med | 74/72 | 18-75 y, office BP $\geq 140$/$>90$ and $<180/110$ mmHg | History of CV or cerebrovascular events, unsuitable renal artery anatomy, eGFR $<40$ ml/min/1.73 m<sup>2</sup> | Change in daytime ambulatory SBP after 2 months | Paradise (ultrasound) | NCT02649426 |
| SPYRAL HTN-OFF MED Pivotal 2020 <sup>16</sup> | Randomized 1:1, RDN vs. sham procedure, off med | 166/165 | 20-80 y, office SBP $>150$ mmHg and $<180$ mmHg, DBP $>90$ mmHg | Angina pectoris, MI within last 3 months, unsuitable renal artery anatomy, eGFR $<45$ ml/min/1.73 m<sup>2</sup> | Change in 24-h SBP after 3 months | Spyral (radiofrequency) | NCT02439749 |
| RADIANCE- | Randomized | 69/67 | 18-75 y, office | History of | Change in | Paradise | NCT02649426 |

| **Study and publication year** | **Study design** | **Sample size (RDN/Sham)** | **Inclusion criteria** | **Exclusion criteria** | **Primary efficacy endpoint** | **Catheter (Modality)** | **Link to the public trial registry** |
| --- | --- | --- | --- | --- | --- | --- | --- |
| HTN TRIO<br>2021<sup>18</sup> | 1:1, RDN vs. sham procedure, on med |  | BP ≥140/>90 mmHg on 3-drug single-pill combination | cerebrovascular or severe CV event, unsuitable renal artery anatomy, eGFR <40 ml/min/1.73 m<sup>2</sup> | daytime ambulatory SBP after 2 months | (ultrasound) |  |
| TARGET BP OFF-MED<br>2022<sup>29</sup> | Randomized 1:1, RDN vs. sham procedure, off med | 50/56 | 18-80 y, office SBP 140-180 and DBP ≥90 mmHg on 0-2 med. | eGFR <45 ml/min/1.73 m<sup>2</sup>, severe HF, unsuitable renal anatomy | Change in 24-h SBP after 2 months | Peregrine (alcohol) | NCT03503773 |
| REQUIRE<br>2022<sup>31</sup> | Randomized 1:1, RDN vs. sham procedure, on med | 72/71 | 20-75 y, resistant hypertension with 3 med., BP >150/>90 mmHg | eGFR <40 ml/min/1.73 m<sup>2</sup>, secondary hypertension, chronic diseases, unsuitable renal artery anatomy | Change in 24-h SBP after 3 months | Paradise (ultrasound) | NCT02918305 |
| RADIANCE II | Randomized | 150/74 | 18-75 y, 24-h | eGFR <40 | Change in | Paradise | NCT03614260 |
| 2023<sup>28</sup> | 2:1, RDN vs. sham procedure, off med | | BP ≥135/85 and $<$170/105 mmHg | ml/min/1.73 m<sup>2</sup>, unsuitable renal artery anatomy | daytime ambulatory SBP after 2 months | (ultrasound) | |
| SPYRAL HTN-ON MED (Pilot and Expansion phase) 2023<sup>30</sup> | Randomized 1:1 for 106 patients and 2:1 for 231 subsequent patients, RDN vs. sham procedure, on med | 206/131 | 20-80 y, 24-h SBP $>$140/90 mmHg on 1-3 drugs | Angina pectoris, MI within last 3 months, unsuitable renal artery anatomy, eGFR $<$45 ml/min/1.73 m<sup>2</sup> | Change in 24-h SBP after 6 months | Spyral (radiofrequency) | NCT02439775 |
| Iberis-HTN 2023<sup>23</sup> | Randomized 1:1, RDN vs. sham procedure, on med | 107/110 | 18-65 y, office SBP 150-180 mmHg and DBP ≥90 mmHg on 3 med | eGFR $<$45 ml/min/1.73m<sup>2</sup>, secondary hypertension, unsuitable renal artery anatomy | Change in 24-h SBP after 6 months | Iberis (radiofrequency) | NCT02901704 |
| Netrod RDN 2023<sup>22</sup> | Randomized 2:1, RDN vs. sham | 139/66 | 18-65 y, office BP ≥ 150/90 and $<$180/110 | eGFR $<$45 ml/min/1.73m<sup>2</sup>, secondary | Change in 24-h SBP at 6 months | Netrod (radiofrequency) | NCT03261375 |
| | procedure, off med | | mmHg on $\geq 2$ drugs | hypertension | | | |
| TARGET BP I 2024<sup>24</sup> | Randomized 1:1, RDN vs. sham procedure, on med | 148/153 | 18-80 y, office SBP $\geq 150$ and $< 180$ mmHg on $\geq 2$ drugs | eGFR $< 45$ ml/min/1.73m<sup>2</sup>, secondary hypertension | Change in 24-h SBP at 3 months | Peregrine (alcohol) | NCT02910414 |

Abbreviations: CV, cardiovascular; DBP, diastolic blood pressure; FMD, fibromuscular dysplasia; HF, heart failure; RDN, renal denervation; med, medications; MI, myocardial infarction; SBP, systolic blood pressure; y, years.

### Effects of renal denervation on ambulatory blood pressure

Independent of the method utilized for renal nerve ablation and compared with a sham procedure, RDN reduced mean 24-hour ambulatory SBP by −4.4 mmHg (95% CI −6.1, −2.7, p<0.00001, Figure 1) as well as 24-hour ambulatory DBP by −2.6 mmHg (95% CI −3.6, −1.5, p<0.00001, Figure 1). There was relevant heterogeneity for both 24-hour ambulatory SBP and DBP (I^2^=68%, and I^2^=60%, respectively). The Iberis-HTN ^23^ and Netrod RDN ^22^ trials were identified as sources of heterogeneity. After excluding these trials, heterogeneity was no longer present (SBP, I^2^=5%; DBP, I^2^=42%) and the 24-hour ambulatory SBP/DBP reductions vs. sham remained statistically significant (mean difference for 24-hour SBP: −3.3 mmHg, 95% CI −4.3, - 2.2, p<0.00001; mean difference for 24-hour DBP: −2.0 mmHg, 95% CI −2.9, −1.0, p<0.0001). Funnel plots for 24-hour SBP and DBP showed no signs of asymmetry (Supplement, Figure 2A and 2B).

**Figure 1.**
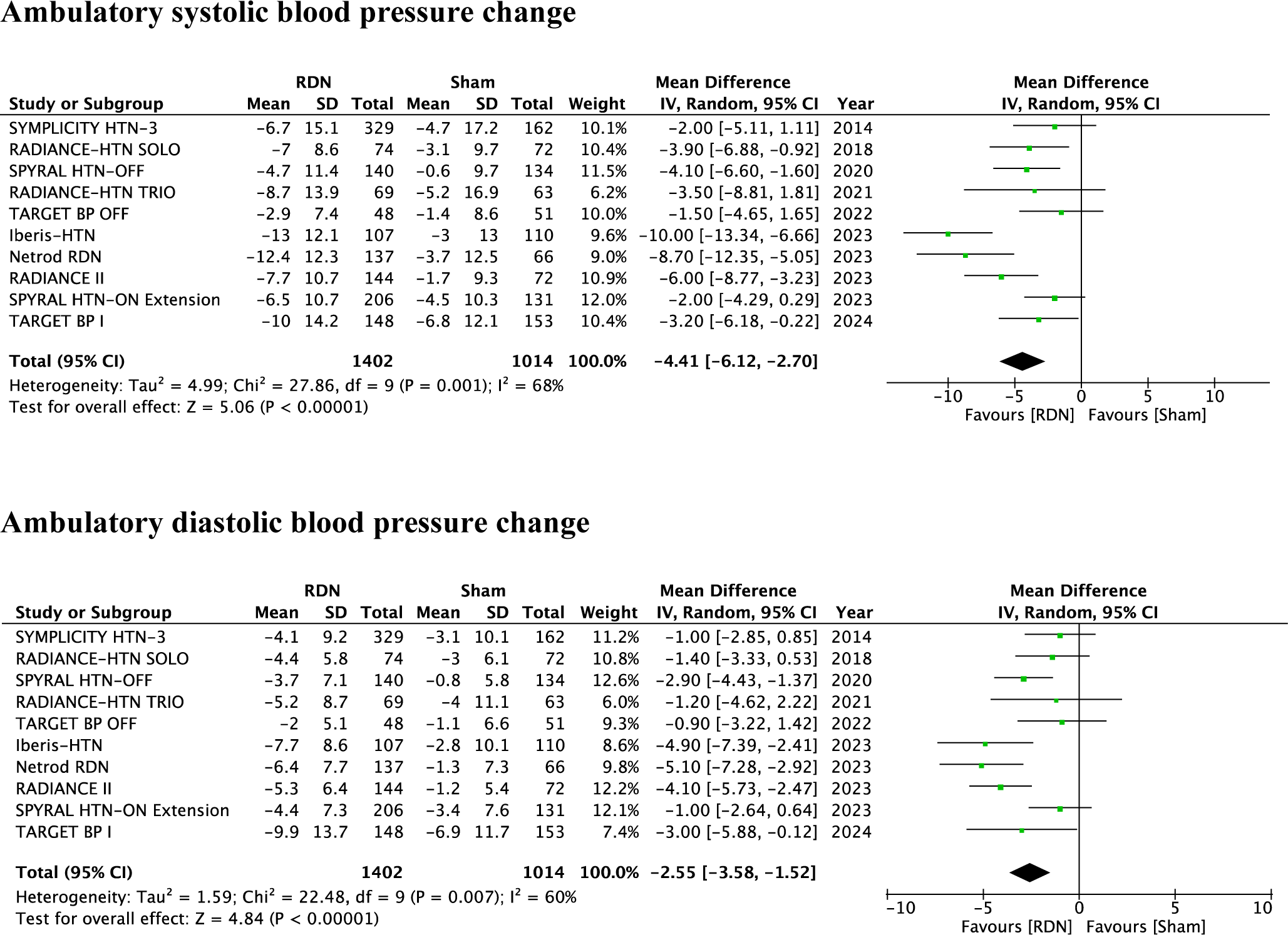
Comparison of effects of renal denervation vs sham procedure on ambulatory systolic (top forest plot) and diastolic (bottom forest plot) blood pressure.

### Effects of renal denervation on office blood pressure

Office SBP and DBP was significantly reduced following RDN (mean difference vs. sham: −6.6 mmHg, 95% CI −9.7, −3.6, p<0.0001 and −3.5 mmHg, 95% CI −5.4, −1.6, p=0.0003, respectively) with relevant heterogeneity (I^2^=82%, Figure 2). The Netrod RDN ^22^ trial was identified as a source of heterogeneity for office SBP and Netrod RDN ^22^ and TARGET BP I ^24^ trials for office DBP. After omitting the data from Netrod RDN ^22^ for SBP and both trials for DBP, the heterogeneity was no longer present with a sustained relevant reduction in systolic and diastolic BP (−5.2 mmHg, 95% CI −6.5, −3.8, p<0.00001, I^2^=0%; −3.1 mmHg, 95% CI −4.0, −2.2, p<0.00001, I^2^=0%, respectively). Funnel plot for office SBP showed presence of asymmetry caused by statistical heterogeneity (Supplement, Figure 3A). The outcomes of the Netrod RDN ^22^ trial were the cause of heterogeneity due the extensive office SBP reductions reported herein. Funnel plot for DBP showed no signs of asymmetry (Supplement, Figure 3B).

**Figure 2.**
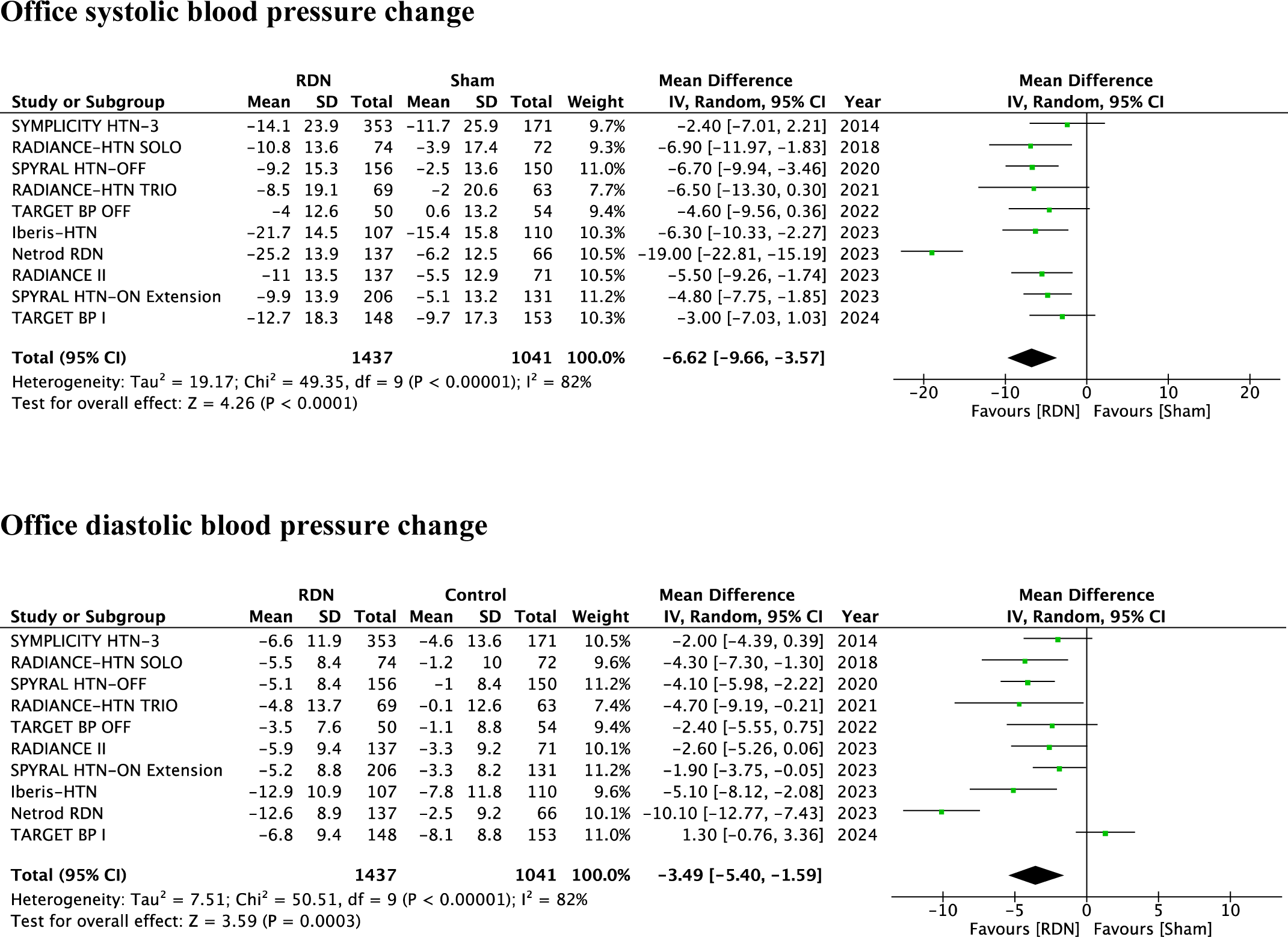
Comparison of effects of renal denervation vs sham procedure on office systolic (top forest plot) and diastolic (bottom forest plot) blood pressure.

### Effects of renal denervation on daytime and nighttime blood pressure

RDN reduced daytime SBP (mean difference vs. sham: −5.2 mmHg, 95% CI −7.6, −2.8, p<0.0001) with relevant heterogeneity (I^2^=76%) and daytime DBP (mean difference vs. sham: - 2.9 mmHg, 95% CI −4.5, −1.3, p=0.0004) with relevant heterogeneity (I^2^=73%; Figure 3). When data from the Iberis-HTN ^23^ and Netrod RDN ^22^ trial were excluded, the relevant heterogeneity for daytime SBP was still present (I^2^=51%) and a significant SBP reduction observed (mean difference vs. sham: −3.6 mmHg, 95% CI −5.5, −1.7, p=0.0002). Nighttime SBP was also significantly reduced by RDN (mean difference vs. sham: −4.5 mmHg, 95% CI −6.1, −2.8, p<0.00001) without relevant heterogeneity (I^2^=32%). Nighttime DBP paralleled these findings (mean difference vs. sham −2.6 mmHg, 95% CI −3.7, −1.5, p<0.00001) without heterogeneity (I^2^=30%; Figure 4).

**Figure 3.**
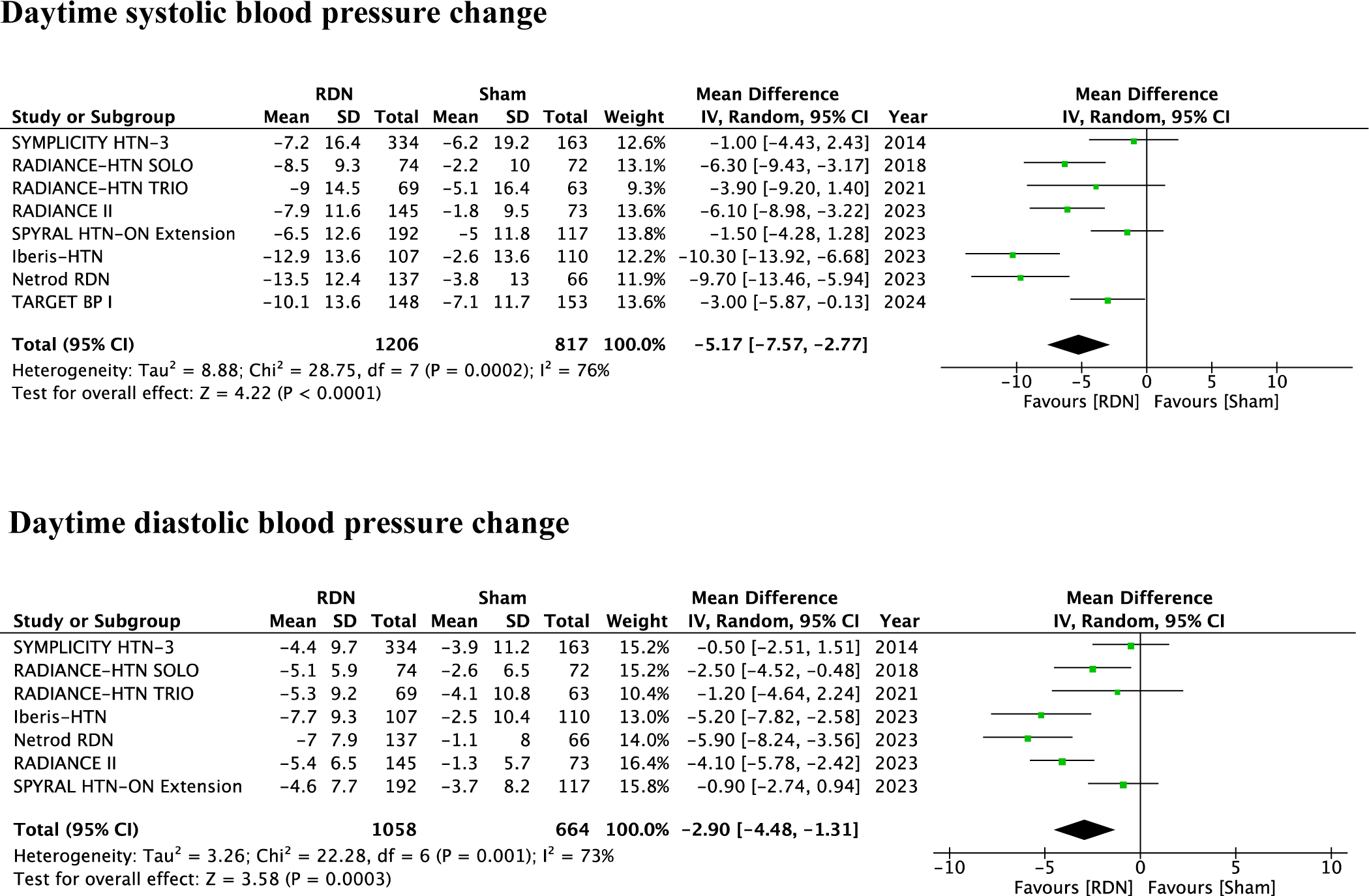
Comparison of effects of renal denervation vs sham procedure on daytime systolic (top forest plot) and diastolic (bottom forest plot) blood pressure.

**Figure 4.**
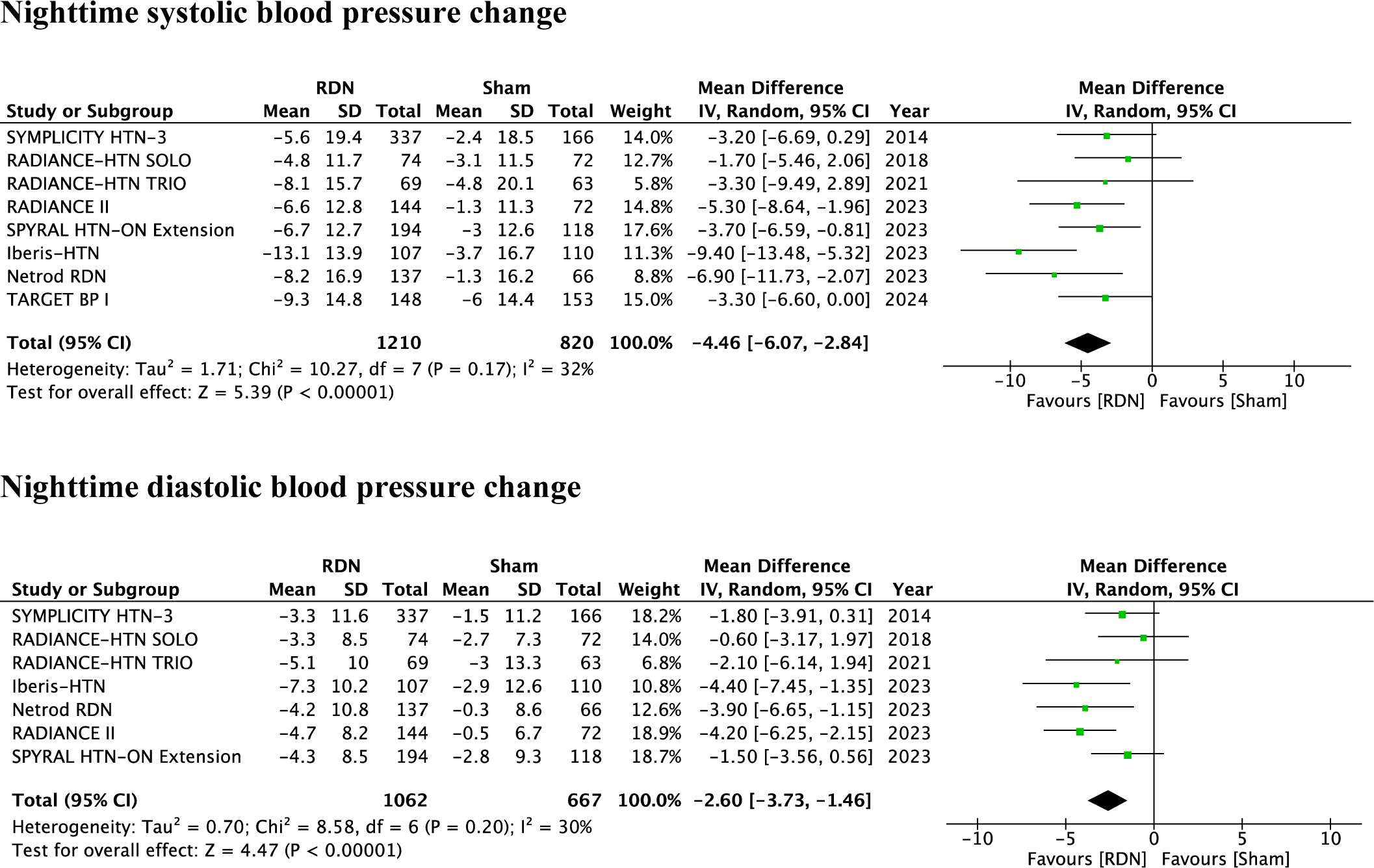
Comparison of effects of renal denervation vs sham procedure on nighttime systolic (top forest plot) and diastolic (bottom forest plot) blood pressure.

### Effects of renal denervation on heart rate

The changes of 24-hour ambulatory heart rate at the time of primary endpoint collection compared with baseline were reported in five trials. ^3, 17, 18, 22, 24, 28^ Heart rate was mildly lowered in the RDN groups (mean difference vs. baseline −2 beats per minute, 95% CI −3.7, −0.2, p=0.03) but not the sham group (mean difference vs. baseline −1.3 beats per minute, 95% CI −3.2, 0.6, p=0.18), without relevant heterogeneity between groups (p for interaction 0.64) (Supplement, Figure 4).

### Sensitivity analysis with long-term data for 24-hour and office SBP

Exploring the long-term data from the SYMPLICITY HTN-3 (36-months of follow up), SPYRAL HTN-ON MED (36-months of follow up), and RADIANCE-HTN and RADIANCE II trials (6-months of follow up) together with other high-quality trials, RDN reduced 24-hour SBP (mean difference −5.4 mmHg, 95% CI −7.8, −3, p<0.00001) and office SBP (mean difference −6.9 mmHg, 95% CI −11.2, −2.6, p=0.001), both with relevant heterogeneity (I^2^=72%, for 24-hour SBP; I^2^=86%, for office SBP) (Supplement, Figure 5). Of note, after the primary outcome assessment, in certain trials, patients were unblinded and medication changes were allowed to achieve BP control.

### Sensitivity analysis with all eligible trials

When performing a sensitivity analysis with all published sham-controlled trials, including those not fulfilling the quality criteria of the ESC/EAPCI clinical consensus statement, RDN reduced 24-hour SBP (mean difference vs. sham: −4.0 mmHg, 95% CI −5.5, −2.4, p<0.00001) and 24-hour DBP (mean difference vs. sham: −2.1 mmHg, 95% CI −3.1, −1.1, p<0.0001), both with relevant heterogeneity (I^2^=62% for SBP; I^2^=60% for DBP) (Supplement, Figure 6). RDN also reduced office SBP (mean difference vs. sham: −6.3 mmHg, 95% CI −9.2, −3.4, p<0.0001) and DBP (mean difference vs. sham: −3.2 mmHg, 95% CI −5.0, −1.4, p=0.0005) with relevant heterogeneity (I^2^=80% for SBP; I^2^=81% for DBP) (Supplement, Figure 7).

RDN reduced daytime SBP (mean difference vs. sham:-4.6 mmHg, 95% CI −6.8, −2.6, p<0.00001) and DBP (mean difference vs. sham: −2.4 mmHg, 95% CI −3.7, −1.0, p=0.0005) with relevant heterogeneity (I^2^=68% for SBP; I^2^=67% for DBP) (Supplement, Figure 8). RDN also reduced nighttime SBP (mean difference vs. sham:-3.5 mmHg, 95% CI −5.4, −1.7, p=0.0001) and DBP (mean difference vs. sham: −1.6 mmHg, 95% CI −3.0, −0.2, p=0.02) with relevant heterogeneity (I^2^=48% for SBP; I^2^=59% for DBP) (Supplement, Figure 9).

### Effects of renal denervation in the absence or presence of antihypertensive medications

There was no relevant difference in 24-hour SBP reduction with RDN vs. sham between trials with or without concomitant antihypertensive medications (−4.8 mmHg, 95% CI −7.7, −2.0, p=0.001, I^2^=79% vs. −4.0 mmHg, 95% CI −5.7, −2.3, p<0.00001, I^2^=32%, respectively; p for interaction 0.62) (Supplement, Figure 10). Similar outcomes were observed for 24-hour DBP reductions (Supplement, Figure 10). There were neither relevant differences in office SBP and DBP reduction between trials with or without concomitant antihypertensive drugs (Supplement, Figure 11) nor were there relevant differences concerning the number of antihypertensive medications prescribed during follow-up (Supplement, Figure 12).

There were more patients in the sham than in the RDN groups with medication adjustments due to safety concerns of meeting escape BP levels (Supplement, Table 16).

### Effects of renal denervation with first-versus second-generation devices

The magnitude of 24-hour SBP and DBP reduction with RDN vs. sham between second-generation versus first-generation RDN devices was not significantly different (SBP: −4.7/-2.0 and DBP: −2.7/-1.0 mmHg, respectively, p for interaction p=0.14 for SBP and p=0.11 for DBP) (Supplement, Figure 13). Similar findings were observed for office SBP (p for interaction 0.10) and DBP (p for interaction 0.30) between second-generation versus first-generation RDN devices (Supplement, Figure 14).

### Effects of radiofrequency-, ultrasound- and alcohol-mediated renal denervation devices

Ambulatory SBP and DBP were significantly reduced in trials investigating RF (mean difference vs sham: −5.2 mmHg, p=0.001 and −2.8 mmHg, p=0.0007, respectively), and ultrasound RDN (mean difference vs. sham: −4.8 mmHg, p<0.00001 and −2.5 mmHg, p=0.02, respectively). Ambulatory SBP was also significantly reduced in alcohol-mediated RDN trials (mean difference −2.4 mmHg, p=0.03), but not DBP (mean difference vs. sham:-1.7 mmHg, p=0.09). There was no relevant interaction between subgroups concerning ambulatory SBP (p for interaction 0.18) and DBP (p for interaction 0.72) (Supplement, Figure 15).

Office SBP was significantly reduced in trials with RF (mean difference vs. sham: −7.9 mmHg, p=0.004), ultrasound (mean difference vs. sham: −6.1 mmHg, p<0.0001) and alcohol-mediated RDN (mean difference vs. sham: −3.6 mmHg, p=0.02), respectively. Similarly, office DBP was significantly reduced in trials with RF (mean difference vs. sham: −4.6 mmHg, p=0.001), ultrasound (mean difference vs. sham: −3.6 mmHg, p=0.0001) but not alcohol-mediated RDN (mean difference vs. sham: −0.4 mmHg, p=0.85), respectively. There was no relevant interaction between subgroups concerning systolic (p for interaction 0.32) and diastolic BP (p for interaction 0.18) (Supplement, Figure 16).

### Safety outcomes

There was no significant difference in the predefined safety outcomes between the RDN and sham groups (Table 2). For all safety outcomes, there were no signs of heterogeneity (I^2^=0%). Furthermore, there was no significant interaction (p for interaction 0.90) regarding change in renal function assessed by change in eGFR from baseline between the RDN (−0.75 ml/min/1.73 m^2^, 95% CI −2.0, 0.5, p=0.24) and sham groups (−0.62 ml/min/1.73 m^2^, 95% −2.2, 1.0, p=0.43) (Supplement, Figure 17).

**Table 2.**
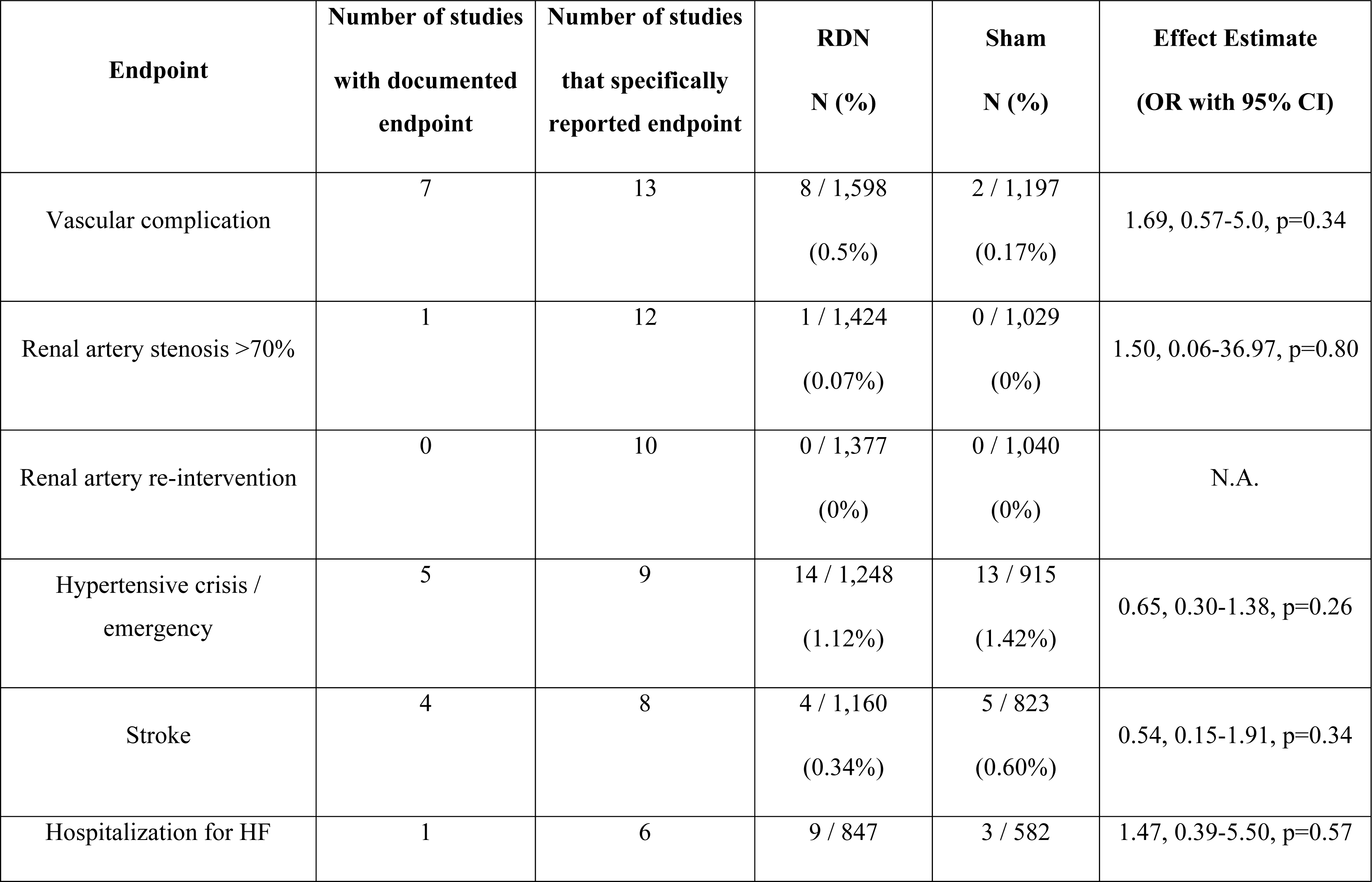
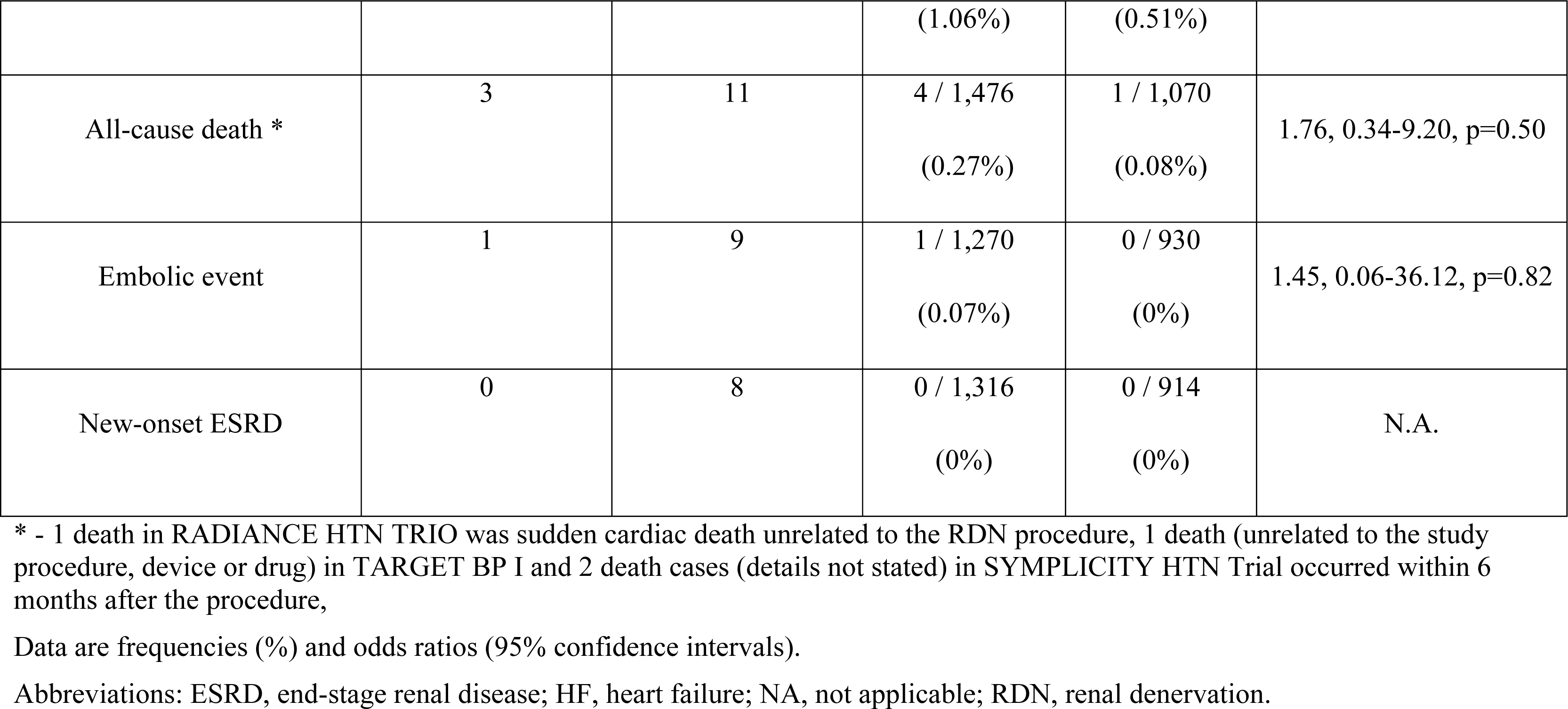
Safety endpoints across RDN and sham group.

## Discussion

In this comprehensive, updated meta-analysis, catheter-based RDN was associated with significantly reduced office and 24-hour BP, including daytime and nighttime SBP and DBP. Subgroup analyses indicated no relevant differences in BP-lowering efficacy regardless of whether individuals were taking or not antihypertensive medication at baseline, or the method or device employed for renal nerve ablation. Moreover, adverse safety events were rare and comparable between the RDN and sham groups.

Hypertension represents the leading modifiable cause of death worldwide. ^36^ The absolute number of people with hypertension is increasing globally and is estimated to reach 1.5 billion by 2025. ^37^ Despite the availability of effective antihypertensive drugs ^38^, guideline-recommended BP targets are often not achieved. ^37, 39^ Device-based antihypertensive treatments have been developed as adjunct treatment options in addition to lifestyle interventions and drugs.

^40^ Several randomized, sham-controlled clinical trials have shown the ability of catheter-based RDN to lower BP in patients with and without antihypertensive medication with mixed outcomes. ^14–18, 28^ Herein, RDN was associated with significantly reduced office and ambulatory BP, including daytime and nighttime BP. The latter has been strongly associated with cardiovascular morbidity and mortality. ^41^

This meta-analysis reports modest but clinically meaningful 24-hour and office SBP reductions of 4.4 mmHg and 6.6 mmHg, respectively. A sensitivity analysis of long-term data suggests that the BP decrease was maintained, if not even continued, following primary outcome assessment after 2-6 months. Indeed, an individual-patient level meta-analysis has shown that a 5 mmHg-reduction in office SBP with antihypertensive drugs reduced the risk for major cardiovascular events (defined as a composite of fatal and non-fatal stroke, fatal or non-fatal myocardial infarction or ischemic heart disease or heart failure causing death or requiring hospital admission) by 10%, regardless of previously diagnosed cardiovascular disease and even in participants with normal or high-normal BP. ^42^

Given the safety and efficacy of RDN, the 2022 ESC Clinical Consensus Statement, the 2023 European Society of Hypertension Guidelines and the 2023 SCAI consensus statement ^43^ recommend that RDN can be considered in patients with resistant hypertension and preserved renal function (eGFR >40 ml/min/1.73 m^2^). ^6, 44^ In line with the 2023 ESH Guidelines for the management of arterial hypertension and the 2022 ESC Clinical Consensus Statements, RDN represents an alternative treatment option for patients with resistant hypertension but also patients with uncontrolled hypertension on fewer drugs if first-line drugs are either not tolerated or patients are non-adherent. ^6^ For most patients, initial antihypertensive therapy should be centered on medications that have been demonstrated to decrease cardiovascular events and mortality. ^6^ Importantly, the decision-making process should involve a multidisciplinary hypertension team and consider the preference of a well-informed patient. Additionally, in the presence of high cardiovascular risk or hypertension-mediated organ damage, RDN should be considered as an additional approach. ^6^ However, it is essential to emphasize that patients must be thoroughly informed by their treating physicians about the potential risks of the invasive procedure. The findings from this meta-analysis support these recommendations since RDN was associated with clinically relevant BP reductions in trials with and without concomitant antihypertensive medication and irrespective of the utilized RDN-modality (RF-, ultrasound-or alcohol-mediated). Thus, these data strongly underpin that RDN as a mechanism earned its unequivocal role in BP control in hypertensive patients.

There was a relevant statistical heterogeneity in several endpoints (24-hour SBP/DBP, office SBP/DBP). According to the sensitivity analysis, the Netrod RDN trial ^22^ alone (for the endpoints office SBP and DBP) and in combination with the Iberis-HTN trial ^23^ (for the endpoint 24-hour SBP) have been identified as sources of heterogeneity due to numerically greater BP reductions following RDN when compared with other trials. One may speculate that this is related to the population included since both trials were conducted in China and studied patients with different background medication, severity of hypertension, and ethnicity. The Iberis-HTN trial, for instance, purposely included younger patients (mean age 45 years) with elevated heart rate (mean of 79 bpm). It has been suggested that RDN may be more effective in younger patients because of a lower incidence of arterial stiffness and an increased sympathetic tone as the leading mechanism of hypertension. ^45^ However, in trials encompassing patients across a wider age spectrum, such as the Radiance HTN trials (which included patients aged 20-75 years) ^34^, sensitivity analyses did not indicate a significant interaction between age and the BP lowering.

Overall, number of vascular complications events was very low (0.5% across all included patients) and did not differ between RDN and sham groups. In a previous meta-analysis, the annual incidence rate of renal artery^34^ stent implantation following RF RDN was 0.2% ^46^, which appears comparable with the natural incidence of renal artery stenosis in patients with arterial hypertension. ^47^ While the analysis suggested a favorable safety profile for RDN, it is crucial to note that the procedures were carried out by experienced physicians, and the data are derived from a relatively small patient cohort.

### Limitations

All included trials excluded patients with known secondary hypertension. Most trials excluded patients with isolated systolic hypertension; thus, limiting generalizability to this hypertension phenotype. However, data from the Global Symplicity Registry ^48^ and a single-center randomized controlled trial comparing different catheter technologies ^49^ suggest that the BP reductions following RDN are not different between patients with isolated systolic hypertension and combined systolic-diastolic hypertension. The maximal follow-up for the assessment of the primary outcome was 6 months. Trials including drug-naïve patients or requiring a washout of antihypertensive drugs had even shorter follow-up durations of 2 to 3 months. The possibility of reinnervation of the renal sympathetic nerves raised the question of whether this would mitigate the effects on BP at the long term. Preclinical studies showed partial, but non-functional, nerve regrowth 30 months after RDN in sheep ^50^ and sustained denervation after 180 days in porcine models. ^51^ Investigating longer-term follow-up is challenging due to several factors. These include the unblinding of patients and outcome assessors, potential crossover of sham patients to RDN, age- and body weight-dependent longitudinal BP changes, the addition of antihypertensive medications to facilitate BP control, and dynamic changes in drug adherence over time. ^6^ In line, the long-term three years results from the Global SYMPLICITY Registry ^52^, the SPYRAL HTN-ON MED^33^, the RADIANCE-HTN SOLO ^53^ and the SYMPLICITY HTN-3 ^32^ trials demonstrated sustained and even progressive effects for at least 3 years. Results of our sensitivity analysis with long-term data of these hallmark trials support these findings. Moreover, this study-level meta-analysis is not based on individual patient data which limits the possibility of performing more detailed subgroup analysis to identify predictors of BP response to RDN.

### Conclusion

This comprehensive meta-analysis of ten randomized sham controlled trials comprising 2,478 patients demonstrated that catheter-based RDN safely reduced 24-hour and office BP up to 6 months compared with sham in hypertensive patients with and without concomitant antihypertensive medication irrespective of the utilized RDN modality.

## Data Availability

All data are available

## Acknowledgements

None.

---

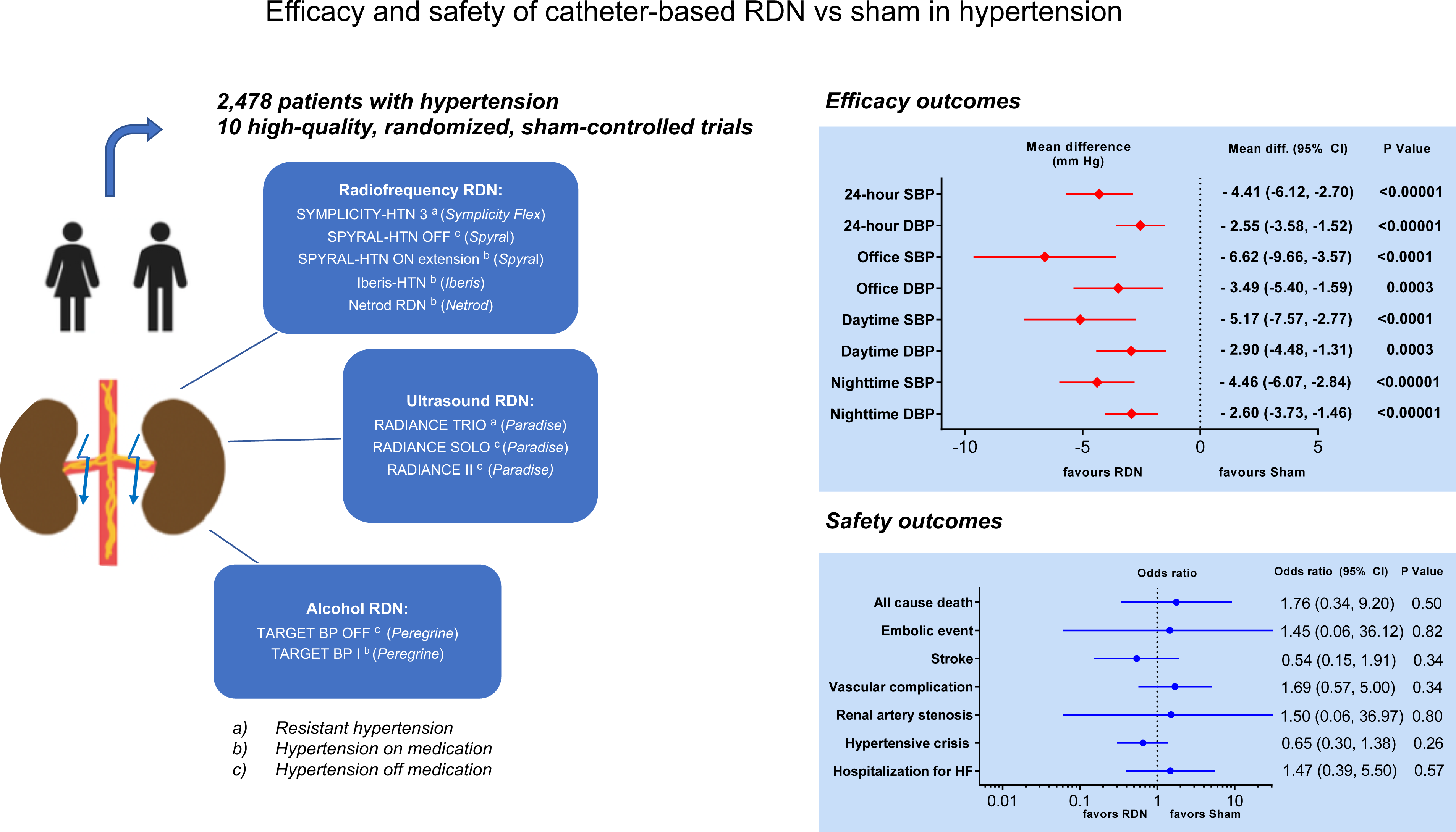

## References

1. DiBona GF. Physiology in perspective: The Wisdom of the Body. Neural control of the kidney. Am J Physiol Regul Integ Comp Physiol. 2005;289:R633–41.

2. Krum H, Schlaich MP, Sobotka PA, Böhm M, Mahfoud F, Rocha-Singh K, Katholi R and Esler MD. Percutaneous renal denervation in patients with treatment-resistant hypertension: final 3-year report of the Symplicity HTN-1 study. Lancet. 2014;383:622–9.

3. Bhatt DL, Kandzari DE, O’Neill WW, D’Agostino R, Flack JM, Katzen BT, Leon MB, Liu M, Mauri L, Negoita M, et al. A controlled trial of renal denervation for resistant hypertension. N Engl J Med. 2014;370:1393–401.

4. Mathiassen ON, Vase H, Bech JN, Christensen KL, Buus NH, Schroeder AP, Lederballe O, Rickers H, Kampmann U, Poulsen PL, et al. Renal denervation in treatment-resistant essential hypertension. A randomized, SHAM-controlled, double-blinded 24-h blood pressure-based trial. J Hypertens. 2016;34:1639-47.

5. Desch S, Okon T, Heinemann D, Kulle K, Röhnert K, Sonnabend M, Petzold M, Müller U, Schuler G, Eitel I, et al. Randomized sham-controlled trial of renal sympathetic denervation in mild resistant hypertension. Hypertension. 2015;65:1202–8.

6. Barbato E, Azizi M, Schmieder RE, Lauder L, Böhm M, Brouwers S, Bruno RM, Dudek D, Kahan T, Kandzari DE, et al. Renal denervation in the management of hypertension in adults. A clinical consensus statement of the ESC Council on Hypertension and the European Association of Percutaneous Cardiovascular Interventions (EAPCI). Eur Heart J. 2023;44:1313–1330.

7. Kandzari DE, Mahfoud F, Weber MA, Townsend R, Parati G, Fisher NDL, Lobo MD, Bloch M, Böhm M, Sharp ASP, et al. Clinical Trial Design Principles and Outcomes Definitions for Device-Based Therapies for Hypertension: A Consensus Document From the Hypertension Academic Research Consortium. Circulation. 2022;145:847–863.

8. Mahfoud F, Azizi M, Ewen S, Pathak A, Ukena C, Blankestijn PJ, Böhm M, Burnier M, Chatellier G, Durand Zaleski I, et al. Proceedings from the 3rd European Clinical Consensus Conference for clinical trials in device-based hypertension therapies. Eur Heart J. 2020;41:1588–1599.

9. Mahfoud F, Schmieder RE, Azizi M, Pathak A, Sievert H, Tsioufis C, Zeller T, Bertog S, Blankestijn PJ, Böhm M, et al. Proceedings from the 2nd European Clinical Consensus Conference for device-based therapies for hypertension: state of the art and considerations for the future. Eur Heart J. 2017;38:3272–3281.

10. Mahfoud F, Böhm M, Azizi M, Pathak A, Durand Zaleski I, Ewen S, Tsioufis K, Andersson B, Blankestijn PJ, Burnier M, et al. Proceedings from the European clinical consensus conference for renal denervation: considerations on future clinical trial design. Eur Heart J. 2015;36:2219–27.

11. Kandzari DE, Bhatt DL, Brar S, Devireddy CM, Esler M, Fahy M, Flack JM, Katzen BT, Lea J, Lee DP, et al. Predictors of blood pressure response in the SYMPLICITY HTN-3 trial. Eur Heart J. 2015;36:219–27.

12. Sakakura K, Ladich E, Cheng Q, Otsuka F, Yahagi K, Fowler DR, Kolodgie FD, Virmani R and Joner M. Anatomic assessment of sympathetic peri-arterial renal nerves in man. J Am Coll Cardiol. 2014;64:635–43.

13. Struthoff H, Lauder L, Hohl M, Hermens A, Tzafriri AR, Edelman ER, Kunz M, Böhm M, Tschernig T and Mahfoud F. Histological examination of renal nerve distribution, density, and function in humans. EuroIntervention. 2023;19:612–620.

14. Townsend RR, Mahfoud F, Kandzari DE, Kario K, Pocock S, Weber MA, Ewen S, Tsioufis K, Tousoulis D, Sharp ASP, et al. Catheter-based renal denervation in patients with uncontrolled hypertension in the absence of antihypertensive medications (SPYRAL HTN-OFF MED): a randomised, sham-controlled, proof-of-concept trial. Lancet. 2017;390:2160–2170.

15. Kandzari DE, Böhm M, Mahfoud F, Townsend RR, Weber MA, Pocock S, Tsioufis K, Tousoulis D, Choi JW, East C, et al. Effect of renal denervation on blood pressure in the presence of antihypertensive drugs: 6-month efficacy and safety results from the SPYRAL HTN-ON MED proof-of-concept randomised trial. Lancet. 2018;391:2346–2355.

16. Böhm M, Kario K, Kandzari DE, Mahfoud F, Weber MA, Schmieder RE, Tsioufis K, Pocock S, Konstantinidis D, Choi JW, et al. Efficacy of catheter-based renal denervation in the absence of antihypertensive medications (SPYRAL HTN-OFF MED Pivotal): a multicentre, randomised, sham-controlled trial. Lancet. 2020;395:1444–1451.

17. Azizi M, Schmieder RE, Mahfoud F, Weber MA, Daemen J, Davies J, Basile J, Kirtane AJ, Wang Y, Lobo MD, et al. Endovascular ultrasound renal denervation to treat hypertension (RADIANCE-HTN SOLO): a multicentre, international, single-blind, randomised, sham-controlled trial. Lancet. 2018;391:2335–2345.

18. Azizi M, Sanghvi K, Saxena M, Gosse P, Reilly JP, Levy T, Rump LC, Persu A, Basile J, Bloch MJ, et al. Ultrasound renal denervation for hypertension resistant to a triple medication pill (RADIANCE-HTN TRIO): a randomised, multicentre, single-blind, sham-controlled trial. Lancet. 2021;397:2476–2486.

19. Stavropoulos K, Patoulias D, Imprialos K, Doumas M, Katsimardou A, Dimitriadis K, Tsioufis C and Papademetriou V. Efficacy and safety of renal denervation for the management of arterial hypertension: A systematic review and meta-analysis of randomized, sham-controlled, catheter-based trials. J Clin Hypertens (Greenwich). 2020;22:572–584.

20. Agasthi P, Shipman J, Arsanjani R, Ashukem M, Girardo ME, Yerasi C, Venepally NR, Fortuin FD and Mookadam F. Renal Denervation for Resistant Hypertension in the contemporary era: A Systematic Review and Meta-analysis. Sci Rep. 2019;9:6200.

21. Page MJ, McKenzie JE, Bossuyt PM, Boutron I, Hoffmann TC, Mulrow CD, Shamseer L, Tetzlaff JM, Akl EA, Brennan SE, et al. The PRISMA 2020 statement: an updated guideline for reporting systematic reviews. BMJ. 2021;372:n71.

22. Zhou Y-J. Effectivness and Safety of the Netrod^TM^ Six-Electrode Radiofrequency Renal Denervation System for the Treatment of Uncontrolled Essential Hypertension. Paper presented at: EURO PCR, May 17, 2023; Paris, France 2023.

23. X Jiang, Rao G. Effectiveness and Safety of The Multi-Electrode Iberis Radiofrequency Renal Denervation System for the treatment of Uncontrolled Primary Hypertension (Iberis-HTN study). Paper presented at: China Interventional Therapeutics (CIT); June 29 - July 2, 2023. Beijing, China.

24. Kandzari DE, Weber MA, Pathak A, Zidar JP, Saxena M, David SW, Schmieder RE, Janas AJ, Langer C, Persu A, et al. Effect of Alcohol-Mediated Renal Denervation on Blood Pressure in the Presence of Antihypertensive Medications: Primary Results from the TARGET BP I Randomized Clinical Trial. Circulation. 2024 Apr 8. Online ahead of print.

25. Sterne JAC, Savović J, Page MJ, Elbers RG, Blencowe NS, Boutron I, Cates CJ, Cheng HY, Corbett MS, Eldridge SM, et al. RoB 2: a revised tool for assessing risk of bias in randomised trials. BMJ. 2019;366:l4898.

26. Deeks JJ, Higgings JPT, Altman DG. Chapter 10: Analysing data and undertaking meta-analyses. In: Higgins JPT, Thomas J, Chandler J, Cumpston M, Li T, Page MJ, Welch VA. Cochrane Handbook for Systematic Reviews of Interventions version 6.4 (updated August 2023). Cochrane, 2023.

27. Altman DG and Bland JM. Interaction revisited: the difference between two estimates. BMJ. 2003;326:219.

28. Azizi M, Saxena M, Wang Y, Jenkins JS, Devireddy C, Rader F, Fisher NDL, Schmieder RE, Mahfoud F, Lindsey J, et al. Endovascular Ultrasound Renal Denervation to Treat Hypertension: The RADIANCE II Randomized Clinical Trial. JAMA. 2023;329:651–661.

29. Pathak A, Rudolph UM, Saxena M, Zeller T, Müller-Ehmsen J, Lipsic E, Schmieder RE, Sievert H, Halbach M, Sharif F, et al. Alcohol-mediated renal denervation in patients with hypertension in the absence of antihypertensive medications. EuroIntervention. 2023;19:602–611.

30. Kandzari DE, Townsend RR, Kazuomi Kario, Felix Mahfoud, Michael A Weber, Roland E Schmieder, Stuart Pocock, Konstantinos Tsioufis, Dimitrios Konstafntinidis, James Choi, et al, on behalf of the SPYRAL HTN-ON MED Investigators*. Safety and Efficacy of Renal Denervation in the Presence of Antihypertensive Medications: SPYRAL HTN-ON MED. J Am Coll Cardiol. 2023;82:1809–1823.

31. Kario K, Yokoi Y, Okamura K, Fujihara M, Ogoyama Y, Yamamoto E, Urata H, Cho JM, Kim CJ, Choi SH, et al. Catheter-based ultrasound renal denervation in patients with resistant hypertension: the randomized, controlled REQUIRE trial. Hypertens Res. 2022;45:221–231.

32. Bhatt DL, Vaduganathan M, Kandzari DE, Leon MB, Rocha-Singh K, Townsend RR, Katzen BT, Oparil S, Brar S, DeBruin V, et al. Long-term outcomes after catheter-based renal artery denervation for resistant hypertension: final follow-up of the randomised SYMPLICITY HTN-3 Trial. Lancet. 2022;400:1405–1416.

33. Mahfoud F, Kandzari DE, Kario K, Townsend RR, Weber MA, Schmieder RE, Tsioufis K, Pocock S, Dimitriadis K, Choi JW, et al. Long-term efficacy and safety of renal denervation in the presence of antihypertensive drugs (SPYRAL HTN-ON MED): a randomised, sham-controlled trial. Lancet. 2022;399:1401–1410.

34. Azizi M, Sharp ASP, Fisher NDL, Weber MA, Lobo MD, Daemen J, Lurz P, Mahfoud F, Schmieder RE, Basile J, et al. Patient-Level Pooled Analysis of Endovascular Ultrasound Renal Denervation or a Sham Procedure 6 Months After Medication Escalation: The RADIANCE Clinical Trial Program. Circulation. 2024;149:747–759.

35. Sterne JA, Sutton AJ, Ioannidis JP, Terrin N, Jones DR, Lau J, Carpenter J, Rücker G, Harbord RM, Schmid CH, et al. Recommendations for examining and interpreting funnel plot asymmetry in meta-analyses of randomised controlled trials. BMJ. 2011;343:d4002.

36. Global burden of 87 risk factors in 204 countries and territories, 1990-2019: a systematic analysis for the Global Burden of Disease Study 2019. Lancet. 2020;396:1223-1249.

37. Worldwide trends in hypertension prevalence and progress in treatment and control from 1990 to 2019: a pooled analysis of 1201 population-representative studies with 104 million participants. Lancet. 2021;398:957-980.

38. Ettehad D, Emdin CA, Kiran A, Anderson SG, Callender T, Emberson J, Chalmers J, Rodgers A and Rahimi K. Blood pressure lowering for prevention of cardiovascular disease and death: a systematic review and meta-analysis. Lancet. 2016;387:957–967.

39. Williams B, Mancia G, Spiering W, Agabiti Rosei E, Azizi M, Burnier M, Clement DL, Coca A, de Simone G, Dominiczak A, et al. 2018 ESC/ESH Guidelines for the management of arterial hypertension. Eur Heart J. 2018;39:3021–3104.

40. Lauder L, Azizi M, Kirtane AJ, Böhm M and Mahfoud F. Device-based therapies for arterial hypertension. Nat Rev Cardiol. 2020;17:614–628.

41. Staplin N, de la Sierra A, Ruilope LM, Emberson JR, Vinyoles E, Gorostidi M, Ruiz-Hurtado G, Segura J, Baigent C and Williams B. Relationship between clinic and ambulatory blood pressure and mortality: an observational cohort study in 59 124 patients. Lancet. 2023;401:2041–2050.

42. Pharmacological blood pressure lowering for primary and secondary prevention of cardiovascular disease across different levels of blood pressure: an individual participant-level data meta-analysis. Lancet. 2021;397:1625-1636.

43. Swaminathan RV, East CA, Feldman DN, Fisher ND, Garasic JM, Giri JS, Kandzari DE, Kirtane AJ, Klein A and Kobayashi T. SCAI position statement on renal denervation for hypertension: patient selection, operator competence, training and techniques, and organizational recommendations. Journal of the Society for Cardiovascular Angiography & Interventions. 2023;2:101121.

44. Mancia G, Kreutz R, Brunström M, Burnier M, Grassi G, Januszewicz A, Muiesan ML, Tsioufis K, Agabiti-Rosei E, Algharably EAE, et al. 2023 ESH Guidelines for the management of arterial hypertension The Task Force for the management of arterial hypertension of the European Society of Hypertension Endorsed by the European Renal Association (ERA) and the International Society of Hypertension (ISH). J Hypertens. 2023;41:1874–2071.

45. Böhm M, Tsioufis K, Kandzari DE, Kario K, Weber MA, Schmieder RE, Townsend RR, Kulenthiran S, Ukena C, Pocock S, et al. Effect of Heart Rate on the Outcome of Renal Denervation in Patients With Uncontrolled Hypertension. J Am Coll Cardiol. 2021;78:1028–1038.

46. Townsend RR, Walton A, Hettrick DA, Hickey GL, Weil J, Sharp ASP, Blankestijn PJ, Böhm M and Mancia G. Review and meta-analysis of renal artery damage following percutaneous renal denervation with radiofrequency renal artery ablation. EuroIntervention. 2020;16:89–96.

47. Chrysochou C and Kalra PA. Epidemiology and natural history of atherosclerotic renovascular disease. Prog Cardiovasc Dis. 2009;52:184–95.

48. Mahfoud F, Mancia G, Schmieder R, Narkiewicz K, Ruilope L, Schlaich M, Whitbourn R, Zirlik A, Zeller T, Stawowy P, et al. Renal Denervation in High-Risk Patients With Hypertension. J Am Coll Cardiol. 2020;75:2879–2888.

49. Fengler K, Rommel KP, Lapusca R, Blazek S, Besler C, Hartung P, von Roeder M, Kresoja KP, Desch S, Thiele H, et al. Renal Denervation in Isolated Systolic Hypertension Using Different Catheter Techniques and Technologies. Hypertension. 2019;74:341–348.

50. Singh RR, McArdle ZM, Iudica M, Easton LK, Booth LC, May CN, Parkington HC, Lombardo P, Head GA, Lambert G, et al. Sustained Decrease in Blood Pressure and Reduced Anatomical and Functional Reinnervation of Renal Nerves in Hypertensive Sheep 30 Months After Catheter-Based Renal Denervation. Hypertension. 2019;73:718–727.

51. Sharp ASP, Tunev S, Schlaich M, Lee DP, Finn AV, Trudel J, Hettrick DA, Mahfoud F and Kandzari DE. Histological evidence supporting the durability of successful radiofrequency renal denervation in a normotensive porcine model. J Hypertens. 2022;40:2068–2075.

52. Mahfoud F, Böhm M, Schmieder R, Narkiewicz K, Ewen S, Ruilope L, Schlaich M, Williams B, Fahy M and Mancia G. Effects of renal denervation on kidney function and long-term outcomes: 3-year follow-up from the Global SYMPLICITY Registry. Eur Heart J. 2019;40:3474–3482.

53. Rader F, Kirtane AJ, Wang Y, Daemen J, Lurz P, Sayer J, Saxena M, Levy T, Scicli AP, Thackeray L, et al. Durability of blood pressure reduction after ultrasound renal denervation: three-year follow-up of the treatment arm of the randomised RADIANCE-HTN SOLO trial. EuroIntervention. 2022;18:e677–e685.

